# HeartBioPortal 3.0: an integrated cardiovascular genomics knowledge environment for molecular, clinical and population-scale interpretation

**DOI:** 10.64898/2026.06.28.26356792

**Authors:** Kasra Vand, Nelson Badia, Bohdan Khomtchouk, Sarath Chandra Janga

## Abstract

Cardiovascular genomics is producing rapidly expanding genetic, molecular, phenotypic and clinical data, yet relevant evidence remains fragmented across resources and difficult to translate into actionable biological and ultimately translational knowledge. HeartBioPortal (HBP) is a browser-based cardiovascular knowledge environment that was developed to address this problem by organizing omics, variant, phenotype and clinical evidence centered around gene queries. Here we describe HBP 3.0, a major update that expands both the data architecture and interpretive interface. This update introduces DataHub, a reproducible data-engineering layer for source ingestion, standardization, variant-centered aggregation, provenance tracking and compact serving artifacts. The release integrates cardiovascular clinical practice guideline context through a graph-backed clinical knowledge layer; incorporates cardiovascular summary statistics from the Million Veteran Program and public aggregate resources; expands source-preserving population frequency, variant annotation and structural-variant; and adds gene profile, drug-discovery and protein-context layers. HBP 3.0 incorporates 594.3 million allele-frequency observations across 18.1 million rsIDs, 3.04 million exon-enriched structural-variant records, 66.9 thousand protein isoforms with 3.26 million non-exon protein feature annotations, 17,128 gene-drug records, and a clinical guideline knowledge graph with 42,895 entities and 106,304 relationships. The redesigned gene dossier view combines phenotype filtering, annotation composition, persistent selected-detail panels and exportable chart data in one workflow. HBP 3.0 is designed to help cardiovascular and eventually cardiometabolic researchers move from a genetic or genomic signal to biological knowledge and potentially clinical and therapeutic context while preserving source provenance and interpretive boundaries. Database URL: https://www.heartbioportal.com/

## INTRODUCTION

Cardiovascular disease (CVD) remains the leading cause of death worldwide (1), motivating large-scale efforts to generate genomic, transcriptomic, phenotypic and clinical datasets for the discovery of disease mechanisms and therapeutic targets. However, the utility of these data is limited by fragmentation across resources, inconsistent data models and the computational expertise required for harmonization and interpretation. As a result, many cardiovascular investigators face a persistent data-to-knowledge gap. In oncology, mature community portals, including cBioPortal and the NCI Genomic Data Commons, have helped transform large-scale cancer molecular profiles and associated clinical attributes into broadly usable research interfaces (2-5). Several cardiovascular resources have begun to address this landscape, including the American Heart Association Precision Medicine Platform, the American Heart Association Protein Atlas Portal and the Cardiovascular Disease Knowledge Portal (6-8). These resources provide important data access, analysis or association-aggregation capabilities, but cardiovascular genomics still lacks comparably integrated, accessible and disease-focused knowledge environments that connect variant, phenotype, population, molecular and clinical context in a unified central resource for benefitting the cardiovascular community.

HeartBioPortal (HBP) was developed to reduce this barrier by transforming heterogeneous cardiovascular omics and genetics resources into browser-based summaries centred around gene and the associated phenotypes. Previous HBP releases integrated genetic association evidence, structural variation, single-nucleotide variants, splicing signals, population allele frequencies and functional annotation layers within a unified query workflow (9-10). The portal builds on the broader cardioinformatics objective of linking bioinformatics infrastructure to precision cardiovascular biology (11,12). Recent usage data indicate increasing engagement with HBP from an international user base, with the strongest activity observed in the United States, China, India, Singapore, the United Kingdom, Germany, and Canada (Supplementary Figure S1). These usage patterns, together with direct feedback from the cardiovascular research community, have identified a need for broader dataset coverage, more responsive visual analytics and outputs that are more directly interpretable in biological, population, clinical and translational contexts.

HBP 3.0 addresses these needs through a set of linked updates to both the data architecture and the user interface. The release introduces DataHub as a reproducible data-engineering layer for source ingestion, standardization, provenance tracking and compact web serving artifacts. It integrates cardiovascular clinical practice guideline context; incorporates cardiovascular summary statistics from the Million Veteran Program (MVP) (13,14); expands source-preserving population frequency context using dbSNP allele-frequency resources (15); adds public summary resources and annotations from TOPMed/dbVar (16-17); expands structural variant (SV) content with a custom visualization module; refreshes variant, association and annotation layers derived from Ensembl, GWAS, ClinVar and dbSNP resources (15, 18, 19) and introduces therapeutic and pharmacologic context through a Drugs & Compounds panel (20, 21). The central goal of this update is not simply to increase data volume, but to connect complementary evidence layers within a single workflow.

HBP 3.0 is intended to complement, rather than replace, large-scale cardiovascular data ecosystems such as TOPMed and BioData Catalyst (16, 22). Controlled-access resources and secure workspaces remain essential for individual-level genomic analysis. HBP addresses a different but related need: it precomputes, harmonizes and serves public or policy-compliant aggregate evidence through a responsive browser interface that helps investigators to inspect cardiovascular genetic signals without repeatedly rebuilding pipelines, reconciling source-specific formats or manually joining disconnected annotation resources. In doing so, HBP 3.0 provides a gene-centered knowledge environment that supports the movement from genetic or genomic signals to biological context, population interpretation and potential clinical or translational relevance.

## HBP 3.0 DATA CONTENT AND IMPLEMENTATION

### Design overview

HBP 3.0 was designed around four linked update pillars that together convert the portal from a collection of cardiovascular association and annotation views into a scalable, gene-centered knowledge environment (Figure 1). First, DataHub provides an open-source data-engineering layer for source ingestion, standardization, quality control, variant-centered aggregation, provenance tracking and publication of compact per-gene and dataset-level serving artifacts. Second, the clinical guideline layer adds graph-backed access to cardiovascular guideline documents, recommendations, supporting excerpts, conditions, therapies, biomarkers and evidence metadata. Third, expanded genomic and contextual evidence layers incorporate large-scale cardiovascular association evidence, source-preserving population allele frequencies, structural-variant resources, gene-profile summaries, protein-context annotations and drug-discovery records from public or license-permitted sources. Fourth, the frontend and visualization system were redesigned around a dynamic gene dossier, with coordinated annotation charts, improved population-frequency views, a new Structural Variant Viewer, an upgraded Protein Consequence Viewer, selected-detail panels, export controls and responsive layout improvements.

**Figure 1.**
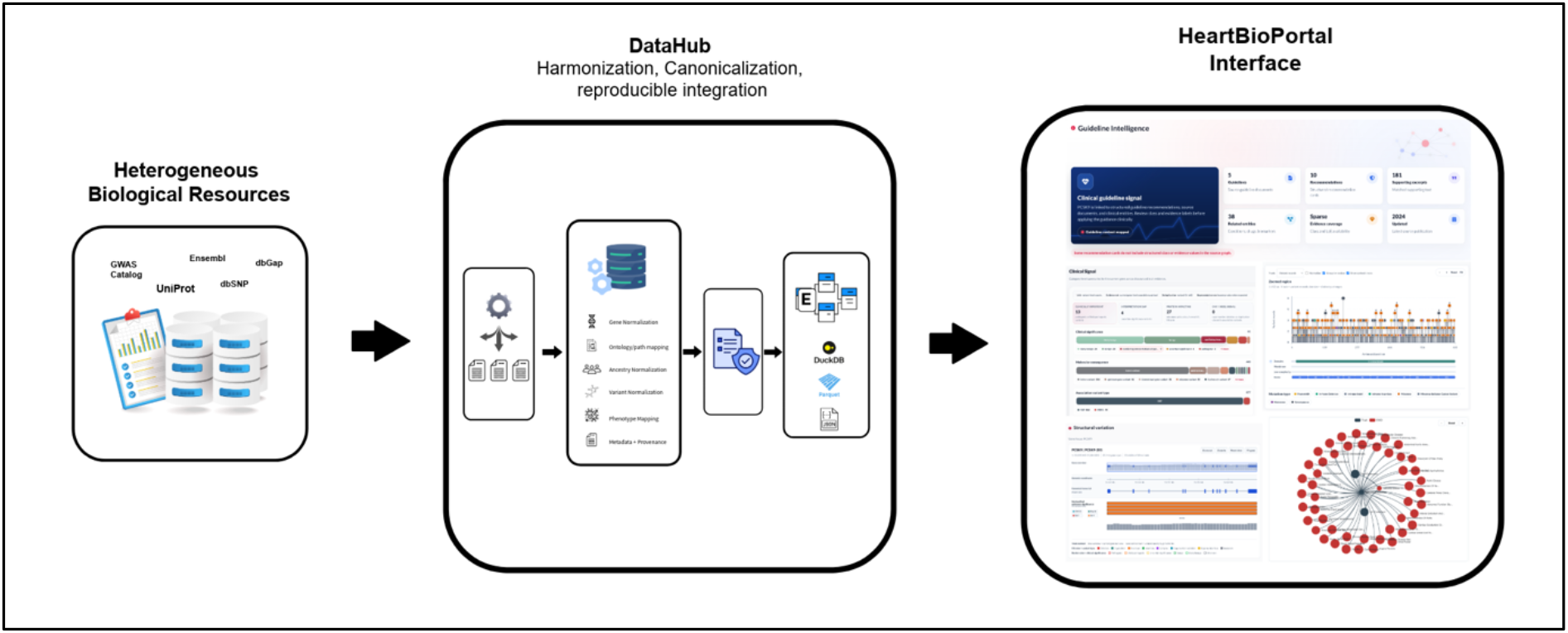
Conceptual architecture of HBP 3.0.

**Figure 2.**
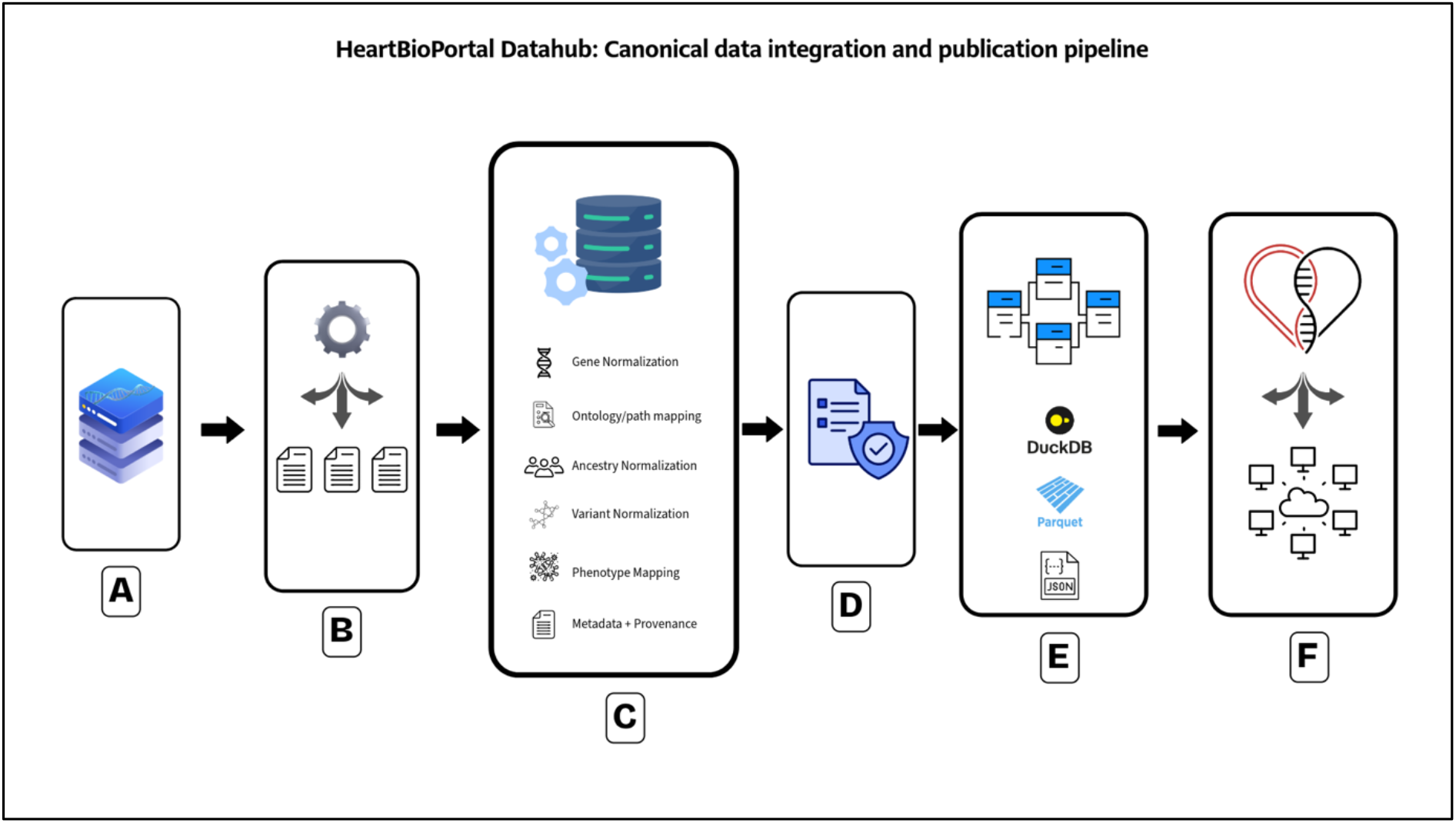
Overview of the DataHub canonical data integration and publication pipeline. (A) Diverse upstream biological data sources, including databases and large-scale consortium resources such as GWAS Catalog, ClinVar, Ensembl, dbSNP, MVP, and TOPMed, enter the pipeline with differing schemas, identifiers, and source-specific formats. (B) Source-specific ingestion and preparation parse these releases, clean raw fields, and standardize input structure into consistent intermediate representations. (C) Canonicalization and semantic harmonization normalize core entities and annotations, including gene and variant identifiers, phenotype mappings, ancestry labels, disease categories, ontology/path assignments, and structured metadata/provenance. (D) Quality control, validation, and enrichment apply dataset contracts and consistency checks to detect malformed records, resolve inconsistencies, and enrich standardized records for downstream use. (E) Standardized evidence is then stored in a unified canonical layer and converted into reproducible downstream artifacts, including reusable canonical records, analytical storage tables (e.g., DuckDB/Parquet), analyzed JSON outputs, serving DuckDB tables, and variant index artifacts. (F) These published outputs are consumed by the HeartBioPortal delivery layer, where backend services and the web application use them to support data querying, visualization, and user-facing knowledge discovery.

### DataHub ingestion, canonicalization and serving architecture

DataHub was introduced as the canonical data-engineering layer for HBP 3.0. It decouples long-running source ingestion, normalization, quality assessment, secondary analysis and artifact publication from the Django runtime that serves the web application. This boundary allows computationally intensive workflows to run in high-performance computing or batch environments, while the web application consumes compact, validated artifacts optimized for search, filtering and visualization. DataHub follows FAIR-oriented design principles by preserving source identity, source version, provenance, schema contracts, licencing metadata and reusable artifact manifests throughout the build process (23).

The canonical association model maps heterogeneous source records into stable HBP chart and API contracts. Each record can retain dataset and source identity, gene and variant identifiers, phenotype or trait labels, disease category, ancestry metadata, statistical evidence, clinical significance and molecular consequence when available, and additional source-specific metadata. Variant identity is treated as a deduplication key for summary visualizations. When the same variant appears in multiple phenotype rows, variant-centered summaries select the strongest association signal as the representative display record, while full detail payloads preserve all source-level evidence. Gene-level charts are recomputed from the complete gene record set rather than by summing phenotype-level summaries, preventing inflation when variants appear in multiple phenotype categories. Snakemake-compatible workflow practices, execution logs and artifact manifests support reproducible regeneration of release-specific HBP data products (24).

### Clinical guideline knowledge graph and API contract

To provide clinically relevant context for gene-centered interpretation, HBP 3.0 incorporates a Heart Clinical Guideline Knowledge Graph (HCG-KG) generated through structured annotation of 42 cardiovascular clinical guidelines from the American College of Cardiology/American Heart Association (ACC/AHA) and the European Society of Cardiology (ESC), obtained from official ACC and ESC guideline sources. In the current release, HCG-KG contains 42,895 imported graph entities/nodes with 9,299 of them being recommendation nodes, and 106,304 edges/relationships with 5,102 of them being class-of-recommendation metadata and 6,860 being level-of-evidence metadata (where reported by the source guideline). HCG-KG links 144 genes to guideline-derived clinical concepts, including associated conditions, diagnostic or prognostic roles, therapeutic implications, biomarkers, interventions, recommendation context and evidence metadata where available. Guideline relationships are represented with source provenance, supporting excerpts, relationship types, recommendation class, level of evidence when reported. HBP serves this layer through a guideline API contract that separates summary responses from detailed records. For a gene query, the summary endpoint returns high-level counts and available clinical-context categories, while detail endpoints provide guideline citations, supporting snippets, recommendation text where permitted, semantic tags, related clinical entities and evidence metadata. In the frontend, these outputs are displayed in a dedicated Clinical Guidelines panel that separates guideline provenance, supporting text and relationship type from the user’s interpretation. This allows users to distinguish formal recommendations from biomarker statements, diagnostic or prognostic context, therapeutic context and indirect gene mentions, rather than treating every guideline occurrence as evidence of clinical actionability.

### Expanded variant annotation and contextual evidence layers

#### 1. Million Veteran Program (MVP)

We integrated publicly available Million Veteran Program (MVP) genome-wide phenome-wide association study summary statistics from dbGaP accession *phs002453*.*v1*.*p1*. This release, titled “MVP Summary Results from Non-Sensitive Omics Studies,” provides aggregate GWAS/PheWAS results from a large VA longitudinal cohort and includes publicly accessible, non-sensitive genomic summary results rather than individual-level participant data (13, 14). The MVP PheWAS resource includes association results across 2,068 traits from up to 635,969 veterans, with results organized by phenotype class, ancestry-stratified analyses, and multi-population meta-analyses. For HBP 3.0, we prioritized cardiovascular and circulatory-system PheCode results and harmonized the association outputs into gene-centered summary layers for interactive exploration alongside other cardiovascular genomic evidence. Within HBP, MVP data are mapped to GRCh38, rsIDs, HGNC gene identifiers and the HBP phenotype tree which is a comprehensive hierarchical record of cardiovascular phenotypes created using public human disease ontology resources (25, 26).

#### 2. Population Allele Frequency

This update also adds a dbSNP-derived allele-frequency datamart comprising approximately 594.3 million source-specific frequency observations across 18.1 million distinct rsIDs (15). Unlike simplified population-frequency displays that collapse all observations into broad ancestry categories, HBP 3.0 retains the original study or resource name, population label, population group, allele label and frequency, sample size, BioProject or BioSample identifiers, genome build and provenance metadata. The resulting layer incorporates frequency observations from 37 studies and resources, including ALFA/Allele Frequency Aggregator, gnomAD v4, 1000 Genomes, 1000 Genomes 30X, TOPMed, PAGE, HGDP-CEPH, HapMap, ExAC, SGDP, 38KJPN and other population-specific cohorts. This design enables users to compare allele-frequency spectra across resources while retaining the context needed to interpret differences in cohort composition, sample size and source ascertainment.

#### 3. Structural Variants

The updated DataHub structural-variant (SV) pipeline integrates dbVar *nstd102*, which contains ClinVar-imported clinically interpreted structural-variant records, together with dbVar *nstd229*, the TOPMed structural-variant call set generated from 138,134 whole-genome sequences (17, 19). DataHub harmonizes SV coordinates, identifiers and gene overlaps to HBP schemas and enriches SV artifacts with Ensembl-derived transcript and exon coordinates (18). The current exon-enriched SV resource contains approximately 3.04 million SV records across approximately 75,000 gene-level payloads. Because a single interval can overlap multiple genes, transcripts or exons, the enriched artifact is intentionally larger than the number of source SV calls and is designed to support interpretable visualization rather than merely reproduce source-level interval files.

#### 4. Protein-context and gene-profile layers

HBP 3.0 adds protein-level context to help users interpret variants beyond genomic coordinates. The Protein Consequence Viewer maps variant and coding-exon evidence onto amino-acid coordinates, allowing users to inspect whether signals fall within annotated isoforms, domains, motifs, regions or other protein features. DataHub builds this layer by combining Ensembl transcript and translation mappings with EBI Proteins and InterPro annotations for protein features, domains, families, motifs and regions (18, 27, 28). The current protein-context resource covers approximately 66.9 thousand isoforms and more than 3.2 million protein feature annotations. This provides a complementary view to the SV viewer which emphasizes genomic intervals, transcript structure and exon overlap, whereas the protein viewer asks where the affected residue or coding region lies in the protein product. HBP 3.0 also adds gene-profile artifacts derived from HGNC, NCBI Gene, UniProt and GOA, providing approved gene nomenclature, stable identifiers, concise biological summaries and structured functional terms for gene-centered interpretation (27, 29–31).

#### 5. Therapeutic and pharmacologic context from drug-discovery resources

HBP 3.0 adds a therapeutic and pharmacologic context layer that supports the Drugs & Compounds panel in the gene dossier. This layer integrates gene-molecule and target-drug relationships from the DrugBank v5.1.12 academic dataset and the Open Targets Platform GraphQL API v4, while preserving source, version, access date and licensing metadata (20, 21). Open Targets records were retrieved through GraphQL queries to the Platform API and harmonized with DrugBank-derived records at the gene, target and molecule levels. For each gene-linked molecule, HBP stores available source-reported metadata including molecule name and type, source provenance, action type, indication, clinical trial phase or trial status where available, mechanism of action, target class, cellular localization, approval or withdrawal status, adverse-event information, chemical classification and pathway annotations. In the current HBP 3.0 build, the merged drug-discovery layer contains 17,128 gene–drug records across 1,839 gene files and 1,454 unique molecule names. These records are intended to provide therapeutic and pharmacologic context for gene-centered interpretation and hypothesis generation.

### Frontend implementation and progressive visualization

HBP 3.0 introduces a redesigned frontend organized into an interface centred around queried gene rather than independent result pages. The interface was updated across the portal to improve readability, responsiveness and interpretive flow, with a consistent visual hierarchy, card-based evidence summaries, clearer typography and spacing, improved pagination, persistent selected-detail panels, unified tooltips, source badges, information icons and chart-level export controls. The visual design was modernized to provide a professional knowledge-portal aesthetic while retaining the HBP red accent theme. Vue-3 components communicate with a store-driven API layer that separates fast summary requests from detail-heavy evidence retrieval, and phenotype filters are applied before aggregation, so charts reflect the selected evidence set rather than hiding elements from an unfiltered display. The visualization layer was rebuilt to support more context-aware interpretation of cardiovascular evidence. Existing annotation, population-frequency, expression and shared phenotype/shared genetic architecture views were upgraded, and a new clinical guideline panel now presents guideline sources, supporting excerpts, recommendation context and related clinical entities. Population-frequency charts were improved to compare source-specific cohorts while preserving sample-size, population-label and provenance information. Two major modules were developed as custom D3.js visualizations: a de novo Structural Variant Viewer and a redesigned Protein Consequence Viewer. Together, these frontend and visualization upgrades make HBP 3.0 more responsive, visually coherent and effective for moving from a gene query to interpretable biological, population and clinical evidence.

## INTEGRATED GENE-CENTERED EVIDENCE DOSSIER IN HBP 3.0

HBP 3.0 brings the new data layers together through a redesigned gene-centered evidence dossier. When a user searches a gene, the portal resolves the query to available DataHub artifacts and returns a compact view of gene identity, biological function, association evidence, variant annotations, clinical guideline context, drug and compound relationships, population-frequency information, structural variation, protein consequence context, expression support and cross-phenotype relationships where available (figure 3). These components are generated as versioned DataHub artifacts with common manifests, compressed per-gene or dataset-level outputs and explicit provenance metadata, allowing the backend to serve new interpretation layers without embedding source-specific logic in the frontend. The result is a stable user workflow in which investigators can move from a candidate gene or association signal to biological, population, clinical and therapeutic context without leaving the portal.

**Figure 3.**
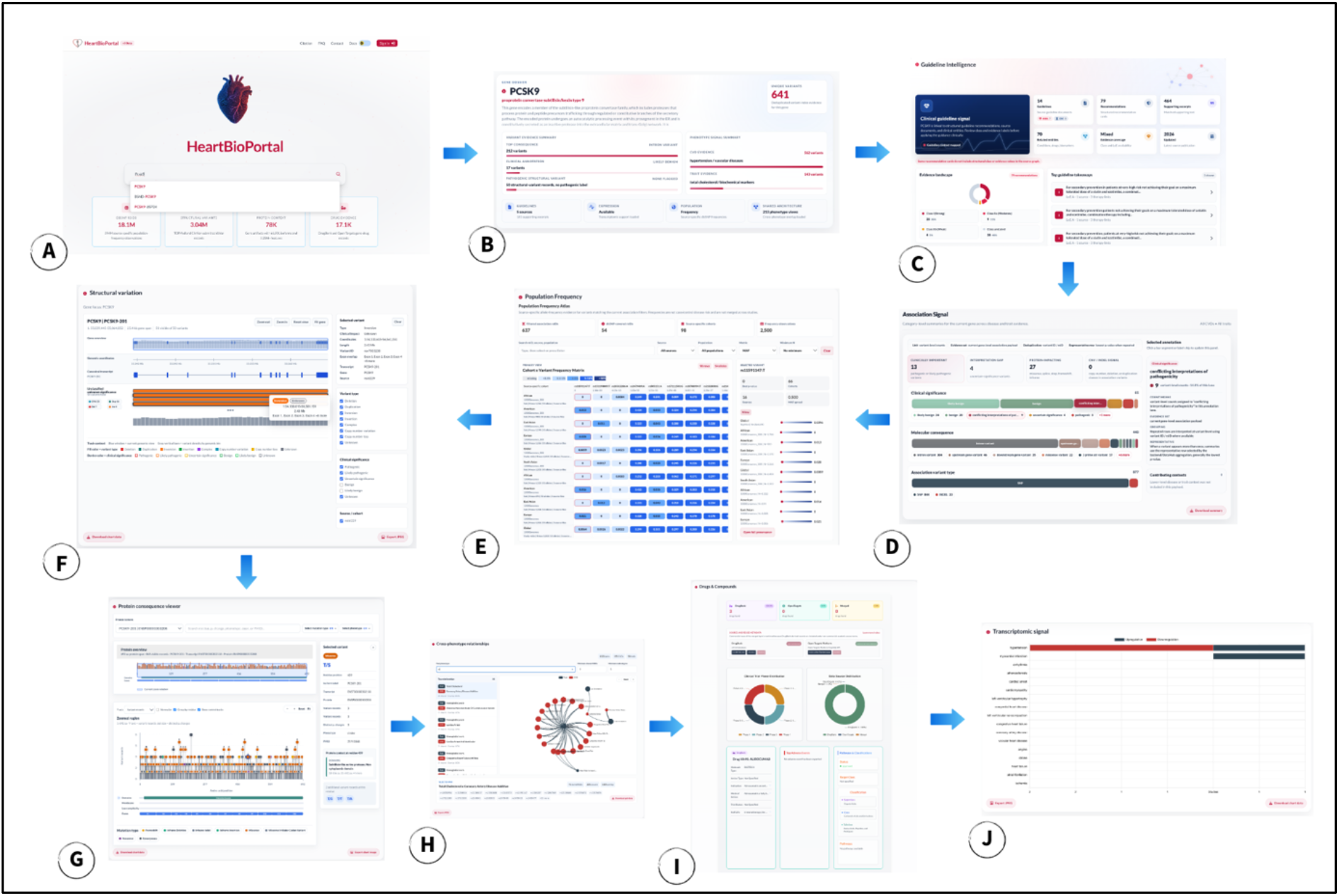
HBP 3.0 gene-centered workflow. Users begin with a gene search (A) and receive an integrated gene dossier summarizing major evidence layers (B). The workflow connects clinical guideline summaries and evidence panels (C) with association signals (D), population frequency (E), structural variation (F), protein consequence mapping (G), cross-phenotype relationships (H), drug/compound annotations (I), and transcriptomic signal (J).

The upper dossier panels summarize association and variant evidence in filterable visual components. Disease and quantitative-trait filters restrict the variant set under inspection, while coordinated annotation charts display variant class, molecular consequence, clinical significance and related categories, preserving missing or unknown values as explicit groups rather than dropping them. The population-frequency view adds source-specific allele-frequency context from the dbSNP-derived datamart, which contains approximately 594.3 million frequency observations across 18.1 million rsIDs from 37 studies and resources. Because these records are joined to association evidence by rsID, displayed frequencies should be interpreted as reference-population context for the selected variants, not as disease-prevalence estimates or association effects. Additional panels provide guideline evidence, drug-discovery context, expression-related signal, shared phenotype relationships, chart exports and source-level provenance; these views are presented as supporting evidence for interpretation and hypothesis generation, not as claims of clinical actionability, therapeutic efficacy or causality.

The lower dossier views support coordinate-aware interpretation of complex variant signals. The SV viewer (Figure 4-A) displays event type, genomic span, clinical significance, exon overlap and source metadata alongside coordinate and transcript tracks. The Protein Consequence Viewer (Figure 4-B) complements this genomic view by mapping variant and exon-level evidence into amino-acid coordinate space which helps users determine whether variants or affected regions intersect protein domains, motifs, regions or other functional features. Together, these visualizations convert HBP search results from separate evidence charts into an integrated cardiovascular genomics interpretation workflow.

**Figure 4.**
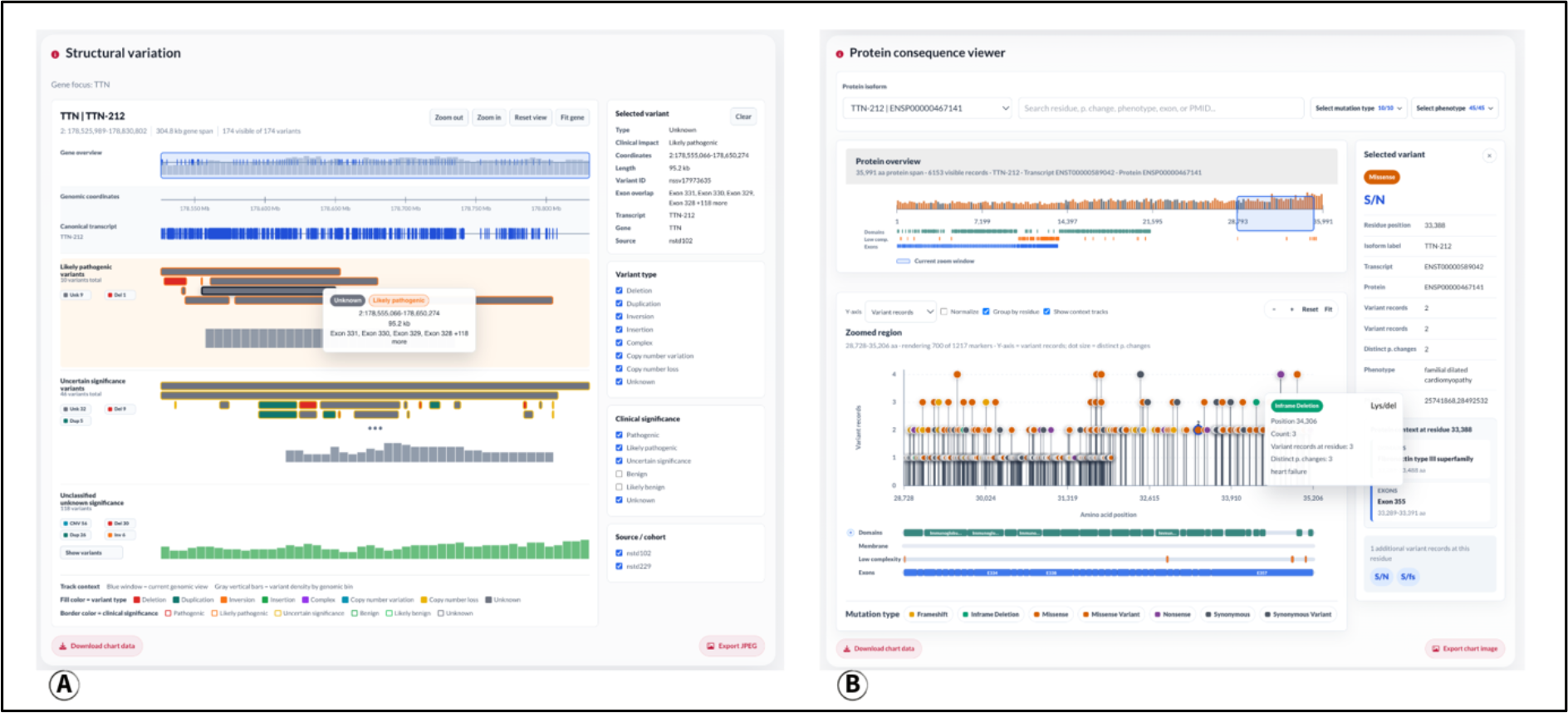
A) Structural variant viewer. B) Protein consequence viewer.

## DISCUSSION

These major upgrades advance HBP from a collection of cardiovascular association and annotation views into a modular, gene-centered interpretation environment. By combining association evidence, source-preserving population frequencies, clinical guideline context, structural-variant exon enrichment, protein-coordinate annotations and gene-profile summaries within a single dossier, the portal allows users to move more directly from a candidate gene or variant signal to its biological, population and potential clinical context. The design is intentionally conservative: HBP does not infer clinical actionability or causality from data co-occurrence alone, and population frequencies, guideline mentions, shared-architecture signals and protein-feature overlaps are presented with source provenance and appropriate interpretive boundaries. However, while HBP 3.0 is a major improvement over previous versions, several limitations remain: Not every gene has complete evidence across all layers, source databases differ in annotation depth and terminology, and controlled individual-level data cannot be redistributed through the public portal. Nevertheless, the DataHub architecture provides a scalable foundation for continued expansion, because new resources can be incorporated as versioned, provenance-aware artifacts without requiring a redesign of the user interface. Future development will extend HBP toward a broader cardiovascular-kidney-metabolic (CKM) knowledge environment, with deeper integration of genetic, transcriptomic, proteomic, clinical, pharmacologic and functional evidence. As these layers mature, HBP will also support more advanced interpretive and translational capabilities, including AI-assisted evidence summarization, variant and gene prioritization, phenotype-aware prediction, and source-grounded knowledge inference, while maintaining transparent provenance and clear boundaries between exploratory hypotheses and clinically actionable conclusions.

## Data Availability

All data produced are available online at https://heartbioportal.com and https://github.com/HeartBioPortal

## DATA AVAILABILITY

HBP 3.0 is freely accessible at https://heartbioportal.com, with documentation and example workflows available at https://heartbioportal.com/docs. Source code, DataHub workflows, source registries, processing configurations and provenance manifests are maintained in the HBP GitHub organization at https://github.com/HeartBioPortal. The manuscript-associated DataHub release, including the DataHub software, source registries, processing configuration files, provenance manifests and build metadata used for the HBP 3.0 evidence layers, has been archived at Zenodo under DOI: https://doi.org/10.5281/zenodo.20277736. This release documents the population-frequency, association, structural-variant, protein-context, gene-profile, expression, shared-architecture and drug-discovery layers described in this manuscript. Clinical guideline extraction resources are maintained in the HCG repository and have been archived at Zenodo under DOI: https://doi.org/10.5281/zenodo.20277758. Clinical guideline knowledge-graph resources are maintained in the HCG-KG repository and have been archived at Zenodo under DOI: https://doi.org/10.5281/zenodo.20277762. Redistributable HBP-derived metadata, manifests and release-associated artifacts are available through the archived GitHub/Zenodo releases and, where applicable, through the HBP download interface. Third-party source data remain subject to their original licenses and access terms. Open Targets-derived annotations were obtained through the Open Targets Platform GraphQL API v4. DrugBank-derived annotations originate from the DrugBank v5.1.12 academic dataset and are subject to CC BY-NC 4.0/non-commercial academic-use terms. Controlled-access individual-level human data are not redistributed through HBP; HBP uses public summary, aggregate or annotation-level resources where permitted and preserves source, version, access-date and provenance metadata in DataHub manifests.

## ACKNOWLEDGEMENTS

The authors thank the HeartBioPortal user community for feedback that informed the HBP 3.0 update. The content is solely the responsibility of the authors and does not necessarily represent the official views of the funding agencies or source-data providers.

## AUTHOR CONTRIBUTIONS

Kasra Vand: conceptualization, data curation, software, methodology, visualization, validation, writing. Nelson Badia: validation, review and editing. Bohdan Khomtchouk: conceptualization, supervision and funding acquisition. Sarath Chandra Janga: supervision, project administration, funding acquisition, review and editing.

## SUPPLEMENTARY DATA

Supplementary Data are available at Database online. Supplementary materials include HBP usage statistics, extended source and artifact manifests, release-level data counts, guideline-source metadata and additional screenshots of supporting evidence panels.

