## Supplementary for "HeartBioPortal 3.0: an integrated cardiovascular genomics knowledge environment for molecular, clinical and population-scale interpretation"

*HeartBioPortal 3.0 supplementary figures, release metrics and source/provenance summaries*

#### Supplementary Data overview

This supplementary file supports the HeartBioPortal (HBP) 3.0 manuscript by providing portal engagement figures, clinical-guideline coverage and interface examples, and release-level tables summarizing major data layers, graph statistics, source provenance and licensing/access notes.

Counts shown here correspond to the HBP 3.0 manuscript-associated DataHub, HCG and HCG-KG releases. Because HBP is updated as source databases change, future portal releases may contain different counts, source versions or artifact coverage. Counts described as payloads or records should not be interpreted as unique human genes unless explicitly stated.

Interpretive boundaries also apply: population frequencies are reference-population context rather than disease-prevalence estimates; guideline links provide clinical context rather than automatic clinical actionability; drug and compound relationships are source-reported therapeutic context rather than evidence of efficacy for a selected cardiovascular phenotype.

### Supplementary figures

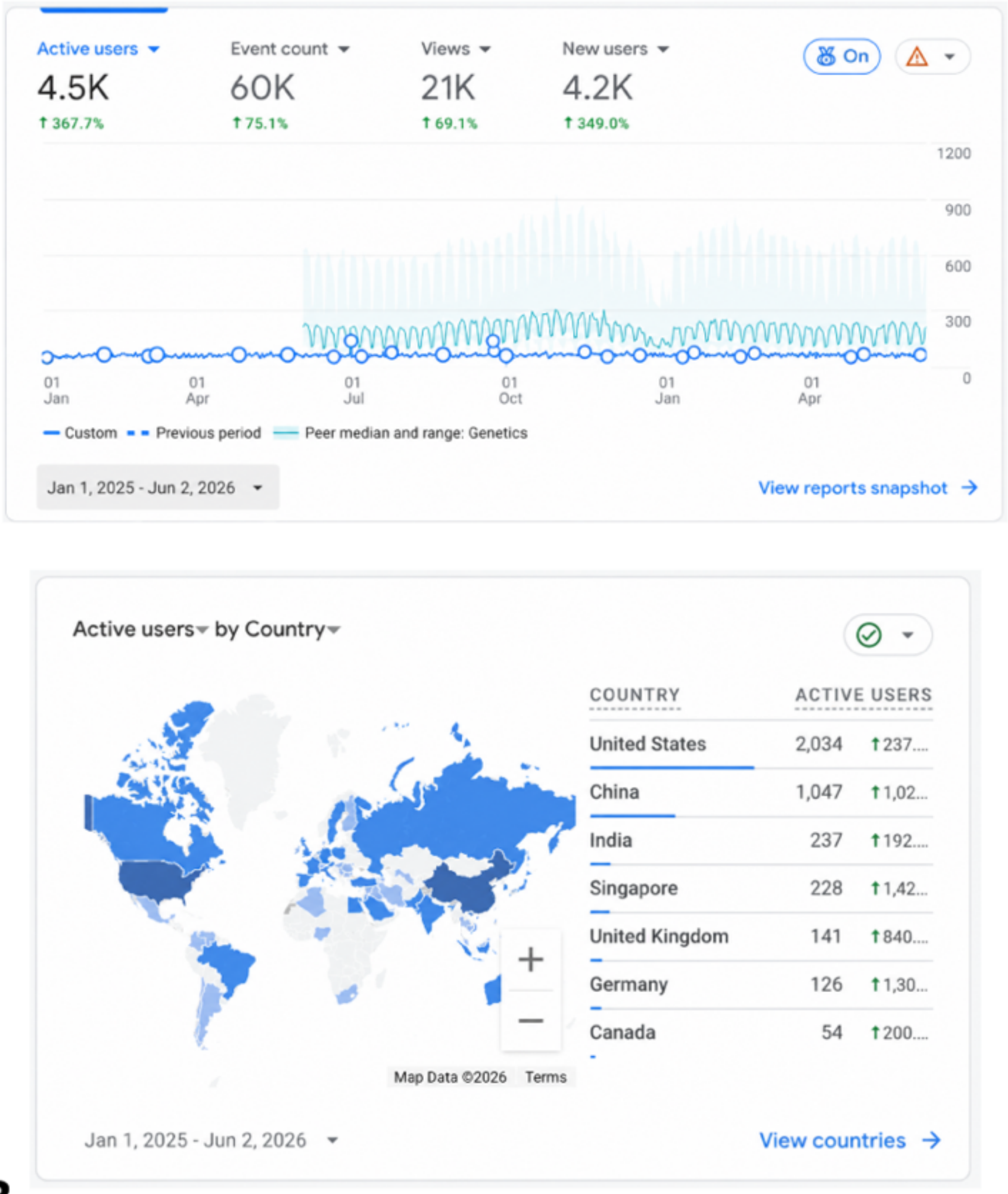


A

B

**Supplementary Figure S1. HBP usage and geographic engagement.** (A) Google Analytics summary for January 1, 2025 to June 2, 2026 showing active users, event count, views, new users and activity over time. (B) Country-level active-user map and table over the same reporting interval. These analytics document portal engagement and geographic reach; they do not measure database completeness or scientific impact.

Alt text: Two-panel screenshot showing HeartBioPortal usage analytics. Panel A displays active users, events, views and new users over time. Panel B displays a world map and table of active users by country.

### Supplementary tables

**Supplementary Table S1. HBP 3.0 release-level evidence layers and user-facing interpretation value.**

| Evidence layer | Implementation in HBP 3.0 | Release-level count or coverage | User-facing interpretation value |
|---|---|---|---|
| DataHub and serving artifacts | Open-source source ingestion, standardization, quality control, variant-centered aggregation, provenance tracking and compact per-gene or dataset-level serving artifacts. | Source registries, processing configurations, manifests and build metadata archived in the HBP DataHub release. | Supports reproducible HBP builds and responsive browser delivery of heterogeneous evidence layers. |
| MVP association evidence | Public Million Veteran Program genome-wide PheWAS summary statistics from dbGaP accession phs002453.v1.p1. | 2,068 traits from up to 635,969 veterans; public aggregate summary results only. | Adds large-scale cardiovascular and circulatory-system association evidence without redistributing individual-level data. |
| Population-frequency context | dbSNP-derived source-preserving allele-frequency datamart including ALFA, gnomAD v4, 1000 Genomes, 1000 Genomes 30X, TOPMed, PAGE, HGDP-CEPH, HapMap, ExAC, SGDP, 38KJPN and other cohorts. | 594.3 million source-specific frequency observations across 18.1 million distinct rsIDs from 37 studies/resources. | Allows users to compare reference-population allele-frequency spectra with source, population, sample-size and provenance context. |
| Structural variants | dbVar nstd102/ClinVar structural variants and TOPMed/dbVar nstd229 call set, harmonized and enriched with transcript/exon context. | Approximately 3.04 million exon-enriched SV records across approximately 75,000 gene-level payloads. | Places interval-based SV evidence in gene, transcript and exon context for gene-centered interpretation. |
| Protein context | Ensembl transcript/translation mappings with EBI Proteins and InterPro feature annotations. | Approximately 66.9 thousand isoforms and more than 3.2 million protein feature annotations. | Supports protein-coordinate interpretation of variants and affected regions in the Protein Consequence Viewer. |
| Gene profiles | HGNC, NCBI Gene, UniProt and GOA-backed gene-profile artifacts. | Available for genes with resolvable source mappings in the HBP build. | Provides concise gene identity, biological synopsis, protein-product and functional-term context. |
| Clinical guideline context | Heart Clinical Guideline Knowledge Graph (HCG-KG) generated from structured cardiovascular guideline annotation. | 42 guideline documents; 62 guideline/source nodes; 42,895 graph entities; 106,304 relationships; 9,299 recommendation nodes; 144 linked genes. | Connects gene searches to guideline documents, recommendations, supporting excerpts, clinical entities and evidence metadata. |
| Drugs and compounds | Merged drug-discovery layer from Open Targets Platform GraphQL API v4 and license-permitted DrugBank v5.1.12 academic dataset. | 17,128 gene-drug records across 1,839 gene files and 1,454 unique molecule names. | Provides therapeutic and pharmacologic context for gene-centered hypothesis generation. |
| Visualization modernization | Redesigned gene dossier, annotation composition, population-frequency charts, clinical guideline panel, Structural Variant Viewer, Protein Consequence Viewer, shared-architecture views and export controls. | New or substantially upgraded UI/visual analytics modules across the HBP 3.0 interface. | Moves users from isolated charts toward coordinated biological, population, clinical and protein-context interpretation. |
| Usage and engagement | Google Analytics reporting interval from January 1, 2025 to June 2, 2026. | Approximately 4.5K active users, 60K events, 21K views and 4.2K new users in the reporting period. | Documents growing community engagement and international usage during HBP 3.0 beta development. |

**Supplementary Table S2. HCG-KG clinical guideline graph release statistics.**

| Metric | Count | Description |
|---|---|---|
| Guideline documents in source corpus | 42 | Structured cardiovascular guideline documents from ACC/AHA and ESC guideline sources represented in the current HCG/HCG-KG release. |
| Guideline/source nodes | 62 | Graph-level guideline or source nodes; this count can exceed the number of source documents because of source grouping, guideline-family representation or versioned source entities. |
| Imported graph entities | 42,895 | Total imported nodes/entities in the manuscript-associated HCG-KG release. |
| Graph relationships/edges | 106,304 | Total relationships connecting guideline entities, recommendations, genes, conditions, therapies, biomarkers, source documents, excerpts and evidence metadata. |
| Recommendation nodes | 9,299 | Structured recommendation entities represented in the graph. |
| Recommendations with class-of-recommendation metadata | 5,102 | Recommendation nodes with class metadata where reported by the source guideline. |
| Recommendations with level-of-evidence metadata | 6,860 | Recommendation nodes with level-of-evidence metadata where reported by the source guideline. |
| Genes linked to guideline-derived clinical concepts | 144 | Genes connected to guideline-derived concepts through structured graph relationships and/or supporting guideline context. |

#### Supplementary Table S3. Key source families, HBP use and access/licensing notes.

| Layer | Source/accession/API | HBP processing/use | Access or redistribution note |
|---|---|---|---|
| MVP association evidence | dbGaP phs002453.v1.p1, MVP Summary Results from Non-Sensitive Omics Studies | Public aggregate GWAS/PheWAS summary statistics are standardized to HBP gene, variant and phenotype models. | Public aggregate results only; no individual-level participant data redistributed. |
| Population frequencies | dbSNP-derived allele-frequency resources and source cohorts including ALFA, gnomAD v4, 1000 Genomes, TOPMed, PAGE, HGDP-CEPH, HapMap, ExAC, SGDP and 38KJPN | Source-specific frequency observations are retained with allele, population label, sample size, study/resource and provenance metadata. | Third-party source terms apply; HBP preserves source and version metadata. |
| Structural variants | dbVar nstd102/ClinVar and dbVar nstd229/TOPMed structural-variant resources | SV intervals are harmonized to HBP schemas and enriched with gene, transcript and exon context for visualization. | Public aggregate/annotation-level resources are used; controlled individual-level sequence data are not redistributed. |
| Protein and gene profiles | HGNC, NCBI Gene, UniProt, GOA, Ensembl, EBI Proteins and InterPro | Identifiers, biological summaries, protein products, transcript/translation mappings, features, domains and functional terms are integrated where available. | Source-specific licenses and attribution requirements apply; provenance is retained in DataHub metadata. |
| Clinical guidelines | Official ACC/AHA and ESC guideline sources; HCG and HCG-KG releases | Guideline-derived recommendations, excerpts, clinical entities and evidence metadata are represented as graph-backed context in HBP. | Source guideline text and excerpts remain subject to original source terms; HBP displays context and provenance, not |

| Layer | Source/accession/API | HBP processing/use | Access or redistribution note |
|---|---|---|---|
| | | | medical advice. |
| Drug-discovery context | Open Targets Platform GraphQL API v4 and DrugBank v5.1.12 academic dataset | Gene-molecule and target-drug relationships are harmonized with molecule type, action, mechanism, indication, trial/status and provenance metadata where available. | Open Targets records follow Open Targets terms; DrugBank-derived records are subject to academic/non-commercial CC BY-NC 4.0 terms and applicable DrugBank licensing. |
| HBP code and build metadata | HeartBioPortal GitHub organization and Zenodo releases | DataHub, HCG and HCG-KG releases provide source registries, processing configurations, manifests and build metadata. | DataHub DOI: 10.5281/zenodo.20277736; HCG DOI: 10.5281/zenodo.20277758; HCG-KG DOI: 10.5281/zenodo.20277762. |

#### Supplementary interpretation notes

- HBP-generated release counts are build-specific. The counts in Supplementary Tables S1-S2 correspond to the manuscript-associated HBP 3.0 release and archived DataHub/HCG/HCG-KG metadata. They may change as source databases are refreshed or additional evidence layers are added.

- The approximately 75,000 structural-variant gene-level payloads should be interpreted as generated payloads or record groupings, not as unique human genes. Exon-enriched SV records may expand beyond the number of source calls because a single interval can overlap multiple genes, transcripts or exons.

- Clinical guideline relationships provide structured context and source provenance. They should not be interpreted as automatic clinical actionability for every gene mention. Class-of-recommendation and level-of-evidence metadata are displayed only where reported by the source guideline and extracted into the HCG-KG release.

- Drug and compound records are source-reported gene-molecule or target-drug relationships. They provide therapeutic and pharmacologic context for hypothesis generation and should not be interpreted as proof of efficacy, regulatory approval or clinical indication for a selected cardiovascular phenotype.

- Population-frequency records are reference-population attributes of selected variants. They are not disease-prevalence estimates and should not be interpreted as association effect sizes.
